# A Stratified Model to Quantify the Effects of Containment Policies on the Spread of COVID-19

**DOI:** 10.1101/2020.04.10.20060681

**Authors:** Vahid S. Bokharaie

**Affiliations:** Vahid Samadi Bokharaie is with Max Planck Institute for Biological Cybernetics, Tübingen, Germany

## Abstract

**Background:** The SARS-Cov-2 virus is spreading fast all over the world and has already imposed a significant social and economic cost in many countries. Different countries have utilised a variety of containment policies with an emphasis on social distancing as a strategy to stop the spread of COVID-19. Quantifying the effects of such policies in different time-scales and comparing the efficacy of various available policies is of utmost importance for policy-makers in different countries. That is even more important when the policy-makers want to plan for long-term strategies and to weigh each option based on its epidemiological effects and socio-economic costs. In the absence of detailed knowledge of the interactions between individuals in a population and the characteristics of the SARS-Cov-2 virus, quantifying the effects of these policies is very difficult. Hence, there is an immediate need for models that can predict the spread of COVID-19 in a population, without the need for such detailed information.

**The Method:** In this manuscript, a method is presented that can be used to predict the spread of COVID-19 in any country and under any containment policy imposed separately on different groups in the population. The method tunes the parameters of a known stratified model based on the available data on the spread of COVID-19. The model includes a set of nonlinear ordinary differential equations and is easy to simulate and easy to understand. As it is presented in this manuscript, the population is divided into age-groups. But given the availability of the data, there is no reason to limit the stratification into only age-groups and we can consider any relevant groups. To estimate the parameters of the model such that it reflects the characteristics of the spread of COVID-19 in a population, the method relies on an optimisation scheme. More specifically, the optimisation scheme estimates the contact rates between different age groups in the population. But a very important and useful feature of the model is that the estimated parameters for one population can be translated and used for any other population with a known age-structure, which in this day and age, includes almost any country or city in the world. Also, it is shown that the method is quite insensitive to the underlying assumptions in the optimisation scheme and also to deliberate or non-deliberate errors that might have occurred in collecting the data.

**Results:** The capabilities of the model are demonstrated using a case study under different containment policies, from an uncontained population to mitigation and suppression policies. Also, we can use the model to quantify the effects of strategies which include switching between different containment policies at pre-determined points in time. The model allows us to quantify the effects of each of these policies on the spread of the disease in a population, in particular, the instantaneous and total numbers of infectious individuals in a population which are important parameters for policy-makers. The simulation results, as explained in the following, provide some insight into various policies and some of the results are possibly counter-intuitive for many people. For example, the model predicts that closing down schools and universities without any other containment policy imposed on older age-groups has almost no impact on the number of patients that might need support-care in hospitals. The method provides an easy-to-use tool to use the information collected on the spread of COVID-19 in a country or region and then use that information in another population which might not have been as widely affected by the disease yet.

## 1 Introduction

There is an abundance of mathematical models of various degrees of sophistication to model the spread of a disease in a population. But there are usually two problems in using these models for real-world applications. One is the difficulty in estimating the values of the parameters of the model based on the characteristics of the disease and the host population. Second is how to translate the requirements of different containment policies to meaningful changes in the parameters of the mathematical model.

There have been some efforts to address both of these issues. The most notable among them for COVID-19 is [9] which is based on long-term effort going back to at least [10]. But those models rely on a detailed knowledge of the population under study which is not easy to replicate for many countries which have not been the subject of similar studies. To fill this gap and to provide an easy to use tool for policy-makers in different countries/regions/cities, in this manuscript, a known stratified epidemiological model is used to predict the spread of COVID-19 in an uncontained population. In the model, the population is divided into different age-groups, as will be explained shortly. Hence, the model can be used to quantify the effects of various containment policies that affect different age-groups differently. The parameters of the model are estimated based on available data on the early stages of the progress of COVID-19 in China. Although the parameters of the model are estimated based on the data collected from China, as we will see in the following, the structure of the model allows us to adapt these parameters for any other country or any population with a known age structure.

## 2 Methods

The mathematical model used for the spread of COVID-19 is an SIR compartmental model, as detailed in SI Section S.1.1. In such models, the population is divided into compartments that capture different stages in the progress of that disease. The three compartments are *Susceptible, S*, which includes those who are healthy and can be infected with the virus; *Infectious, I*, which includes those who are infected and can transmit the disease; and *Removed, R*, which includes those who were Infectious but are not any more, because they recovered or because they died. This choice is guided by the common consensus among immunologists that people have short-term immunity to seasonal coronaviruses. For the SARS-CoV-2 virus, it is already shown to be the case for Macaque monkeys in [5] and for humans in [4]. Although there has been a report of a patient recovered from COVID-19 to be infected again [11], which if proved to be a common phenomenon, then SIS models would be more suitable for COVID-19. But in the absence of overwhelming evidence to the contrary, assuming short-term immunity for COVID-19 seems to be a reasonable choice. Although in the interest of completeness, the mathematical descriptions for both SIS and SIR models are presented in Section S.1.

One important feature of the model used in this manuscript is that in the model, each compartment is divided into groups. The choice of how to group the population is completely arbitrary from a theoretical perspective. But from a practical point of view, such a choice should be informed by the characteristics of the disease and also the availability of the relevant data used to tune the parameters of the model. And in that regard, a very good choice for modelling the spread of COVID-19 is to divide the population into age groups. More specifically, the population in each compartment *S, I* and *R* is divided into nine age groups 0-10, 10-20, …, 70-80 and 80+. And the subsequent model is a set of 3×9 nonlinear ordinary differential equations (ODEs) with some unknown parameters which we should estimate based on the known characteristics of the SARS-CoV-2 virus. These parameters are *transfer rates*, and *contact rates*. Transfer rate, represented with *γ*_*i*_ for each of the age-groups *i* = 1, …, 9, is the rate at which individuals leave the Infectious compartment and is relatively easy to estimate. It depends on the average time period between the moment an individual becomes Infectious and the moment that the individuals are considered cured or die. There are some estimates for this time period, as reported in [2]. I have assumed this time-period for COVID-19 to be 20 days. Considering one day to be the unit of time, and assuming this time-period is on average the same for all age groups, then *γ*_*i*_ = 0.05 for *i* = 1, …, 9.

Another set of parameters are contact rates, represented with *β*_*ij*_ for *i, j* = 1, …, 9 which denote the rate at which Susceptibles in age-group *i* are infected by Infectious individuals in age-group *j*. Contact rates are very difficult to estimate because they capture various characteristics of the population and the dynamics of the virus. Their value can depend on the average number of direct contacts between the members of different groups, which needs a comprehensive and detailed analysis of the behaviour and mobility of the individuals in a population. Contact rates can also depend on the differences in susceptibility of each age-group to the virus and also on the mechanisms of its transmission. Hence, to directly calculate the values of contact rates is a monumental task that even if possible, might be extremely difficult and time-consuming.

In this manuscript, a method is presented that allows us to overcome the difficulties in estimating contact rates between individuals or groups in a population, and estimate these parameters indirectly based on the available data. The method uses an optimisation scheme to achieve this goal. The optimisation scheme is based on two distinct but important sets of data on the spread of COVID-19. One is the estimate for basic reproduction number, *R*_0_, for COVID-19 in the early stages of the spread of the virus. There are various estimates for that parameter, but the one reported in [15], which is *R*_0_ = 2.28 is used in this manuscript. A few other groups have also reported values very close to that [2]. The other piece of information is coming from [1], which shows the relative distribution of confirmed cases of COVID-19 in each age-group in China as of February 11th, as shown in SI Section S.2. Up until two weeks before this date, Chinese authorities did not impose any meaningful containment strategies and the virus was spreading in an uncontained population. Hence we can assume to most part these numbers are the results of the uncontained spread of the virus. But what makes the adopted optimisation scheme so effective is that it relies on the ratio of confirmed cases in age-groups, not their absolute values. The advantage of this choice is that it makes the resulting model quite insensitive to the underlying assumptions in the optimisation scheme. The other advantage is that relying on the absolute numbers of confirmed cases can make the methodology vulnerable to deliberate or non-deliberate errors that might have occurred in the reported data. But assuming similar probabilities of occurrences of such errors in different age-groups, the ratio between these values would be less affected.

Mathematical details of both the model and the optimisation schemes are explained in detail in the Supplementary Information section. The important thing to keep in mind is that although the optimisation scheme uses data collected from China, the structure of the mathematical model is such that it allows us to use the obtained values for contact rates in any other population or country. The model has some advantages and disadvantages which are discussed in Section 4. But it can serve as a useful first step to evaluate various containment policies in different countries. Next section explains some of these applications.

## 3 Results

Using the methodology explained in the previous section, we can predict the trajectory of Infectious and Removed compartments in each age group in any population with a known age-structure (which includes almost every country in the world). In the following, we can see results of such simulations for different countries, with an emphasis on Iran. Iran has been chosen as a case study mainly because among the countries which are affected sooner and wider, has been the one with almost no effective containment strategy in place, and such a policy can lead to consequences as explained in the next section.

### 3.1 Uncontained Spread of the Virus

Let’s consider the case in which COVID-19 is spread uncontained, i.e. the case that people in the society interact with each other as in normal times, with no external or self-imposed restrictions in the interactions.

Figure 1.1shows how the ratio of Infectious people changes in the 18 months after the introduction of SARS-CoV-2 virus in the population. And Figure 1.2shows the trajectory of the Removed ratio in each population. As a reminder, Removed compartment includes those who were Infectious, and then were cured or died. It should be noted that the model is not directly concerned with the mortality rate or the number of those who might need respiratory or intensive care in hospitals. Such information can be inferred from the number of Infectious and Removed compartments in each country, based on the available data or estimates.

**Figure 1.1:**
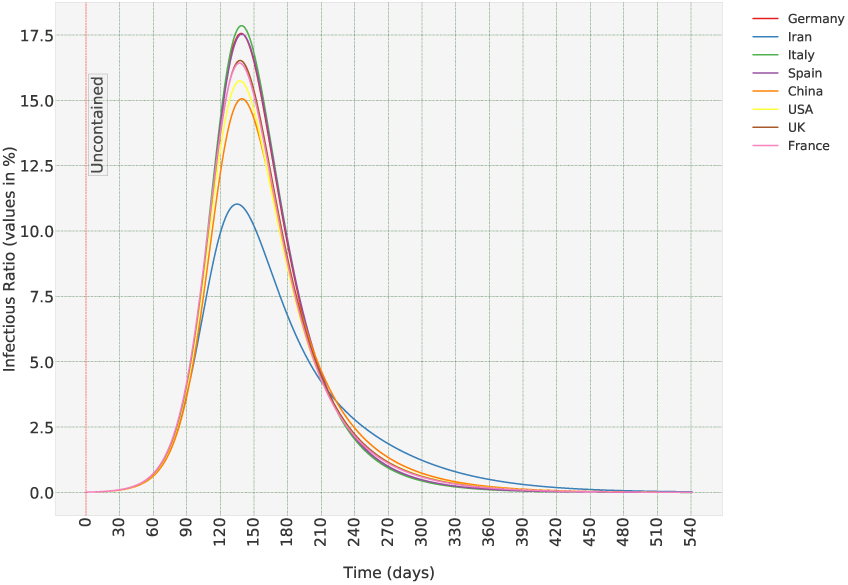
Aggregate ratio of Infectious in the uncontained scenario in different countries.

**Figure 1.2:**
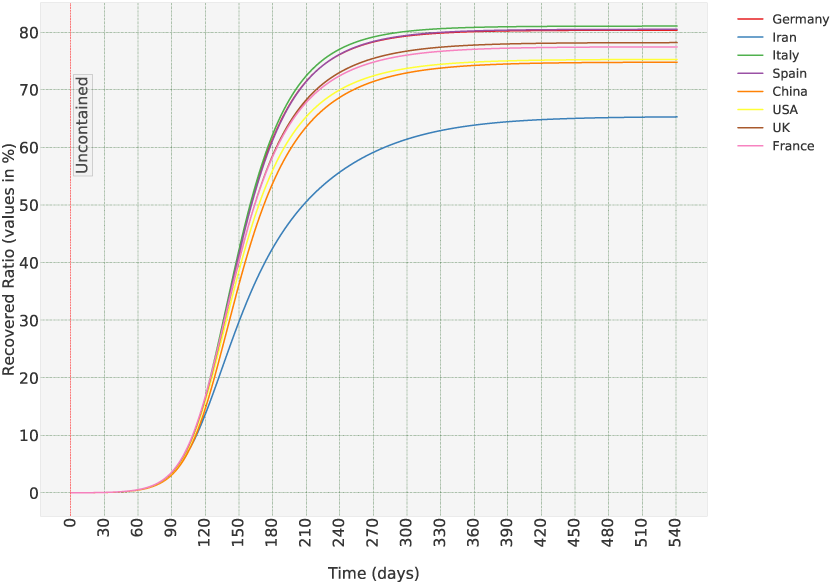
Aggregate ratio of Removed compartment in the un-contained scenario in different countries.

As can be seen, the model predicts the peak in the number of Infectious people and the eventual ratio of the Removed population which varies considerably among different countries. That can be explained based on differences in the population pyramids in these countries. For example, in Iran, 66.8% of the population are under 40, while in Italy only 39.8% are under 40 years old [3]. The model predicts that countries with an older population would be affected more if they let the virus spread uncontained, in agreement with for example [9] when comparing mortality rates in the USA and UK.

To solve any system of Ordinary Differential Equations (ODEs), apart from the equations themselves, we should also define the initial conditions, which in our case means the initial ratios of Infectious and Removed populations in each age group. In all the figures presented in this manuscript, I have assumed 1 in 10,000 in each age group is Infectious, and the Removed population is 0 in Day 1. For a country of 10 million, that amounts to 1,000 Infectious in total. We should be cautious in using a model based on a system of continuous ODEs for numbers lower than that. It should be noted that the peak values in Figures 1.1 and 1.2 are barely affected by the choice of initial conditions. But that is not true for the time it takes to reach the peak values. Hence, in order to predict the day in which the number of Infectious reaches the peak value, we should have a reasonable estimate of the initial conditions. But given the fact that usually in early stages of the spread of the virus, numbers usually remain hidden from authorities, initial conditions are almost impossible to guess. But the good news is that having a good estimate of the numbers of Infectious and Removed ratios in the age groups, at any moment in time, not necessarily the early stages of the spread of the disease, allows us to find the initial conditions that will result in that specific solution in that specific instance in time, and from there, we can predict the time to reach the peak value of the Infectious ratio.

Figures 1.3 and 1.4 show the Infectious and Removed population in each age-group in Iran. As mentioned in the previous section, the output of the model is the trajectory of the spread of the disease in each age group, and the plots in Figures 1.1 and 1.2 are calculated using the relative distribution of the population of each country in one the nine age-groups; 0-10, 10-20, …, 60-70, 70-80, 80+. The same applies to any aggregate plot presented in this manuscript. This characteristic of the model is particularly useful when we want to evaluate the effects of different containment strategies applied to different age groups, as shown in the following.

**Figure 1.3:**
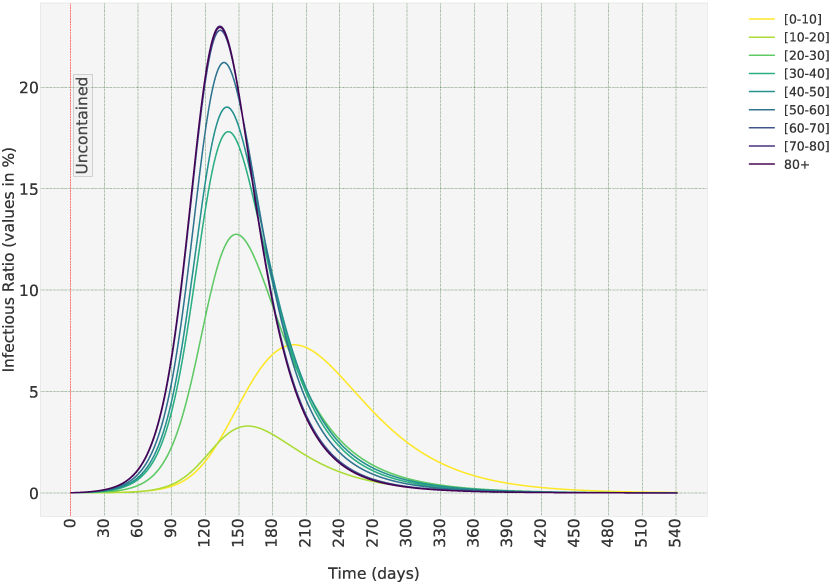
The Infectious ratio in the uncontained scenario in each age-group Iran.

**Figure 1.4:**
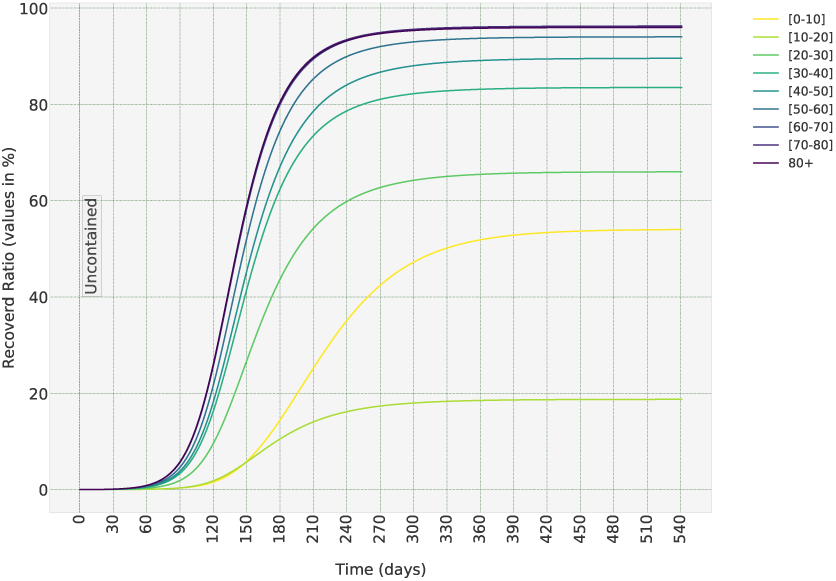
The Removed ratio (ratio of those who were infected and then were cured or lost their lives) in the uncontained scenario in each age-group Iran.

### 3.2 Suppression Strategies

The advantage of having a stratified model is that we can quantitatively evaluate the effects of various containment strategies that affect different age-groups differently. Table 2 has listed some of the more common policies and how they are defined. The column under *Policy Description* defines to what extent the contacts of some age-groups are assumed to be decreased under each policy. These values are chosen intuitively, but any other definitions and any other policies can be easily defined in the software package that is developed as a part of this study [6].

**Table 1:** Maximum Instantaneous Infectious ratio and eventual Removed ratio in different countries in the uncontained scenario. The Removed compartment includes those individuals who have been Infectious and then were cured or died.

| Country | Eventual Ratio of Removed Population (%) | Maximum Instantaneous Infectious Ratio (%) |
| --- | --- | --- |
| Germany | 80.34% | 17.56% |
| Iran | 65.29% | 11.03% |
| Italy | 81.05% | 17.86% |
| Spain | 80.49% | 17.54% |
| China | 74.78% | 15.06% |
| USA | 75.27% | 15.75% |
| UK | 78.18% | 16.53% |
| France | 77.44% | 16.43% |

**Table 2:** Effects of different policies in Iran, when COVID-19 is spread uncontained for the first 90 days, assuming 1 in 10,000 in the population are Infectious on Day 1.

| Policy Label | Policy Name | Policy Description | Eventual Removed Ratio (%) | Maximum Instantaneous Infectious Ratio (%) | $R_0$ |
| --- | --- | --- | --- | --- | --- |
| UN | Uncontained | All interactions as in normal circumstances | 65.29 | 11.03 | 2.28 |
| KI | Only Schools and Universities Closed | interactions of [0-20] age groups decreases to 20% | 47.20 | 9.80 | 2.21 |
| KIOF | Schools, Offices and Companies Closed | Combination of KI and interactions of [20-70] age range decreases to 50% | 24.97 | 3.98 | 1.49 |
| EL | Only Elderly Social Distancing | interactions of 70+ age groups decreases to 25% | 63.36 | 8.55 | 1.79 |
| KIEL | Schools Closed Elderly Social Distancing | combination of KI and EL | 39.08 | 5.60 | 1.68 |
| SD | Social Distancing | combination of KI and EL and interactions of people in [20-70] age range reduce to 20% | 9.46 | 3.72 | 0.49 |
| LD | Lock-down | interactions of all individuals reduced to 10% | 7.90 | 3.72 | 0.23 |

As can be seen in Figures 2.1 and 2.2, all the containment policies decrease both the peak instantaneous Infectious ratio and eventual Removed ratio, which is to be expected. The most effective policy among the ones listed in the Table 2 is *Lockdown*. Although it does not differ much from SD, in which individuals in the population have 20% or 25% interactions compared to normal times, compared to only 10% in Lock-down.Depending on the social and economic circumstances in the country, the slight decrease in peak Infectious ratios might not be worth the cost of imposing a much more invasive policy such as total lock-down.

**Figure 2.1:**
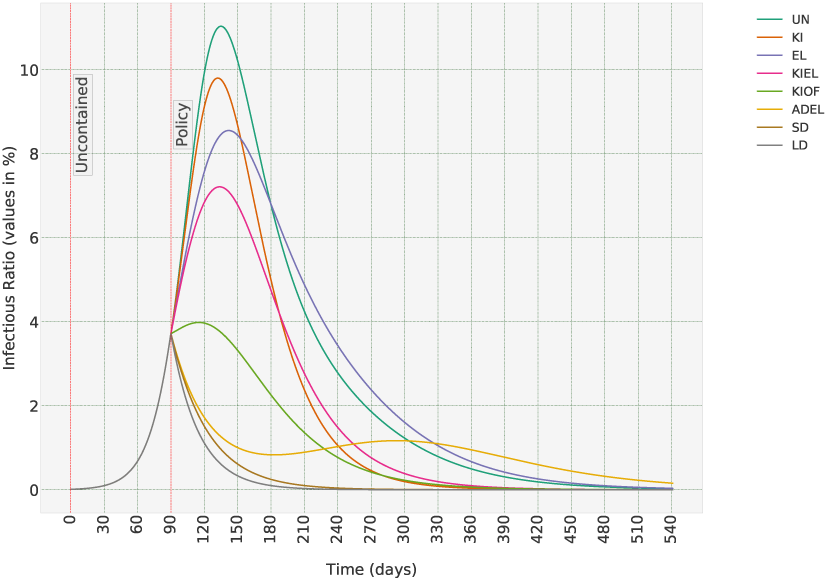
How Infectious ratios change under different containment policies. Policies are defined in Table 2

**Figure 2.2:**
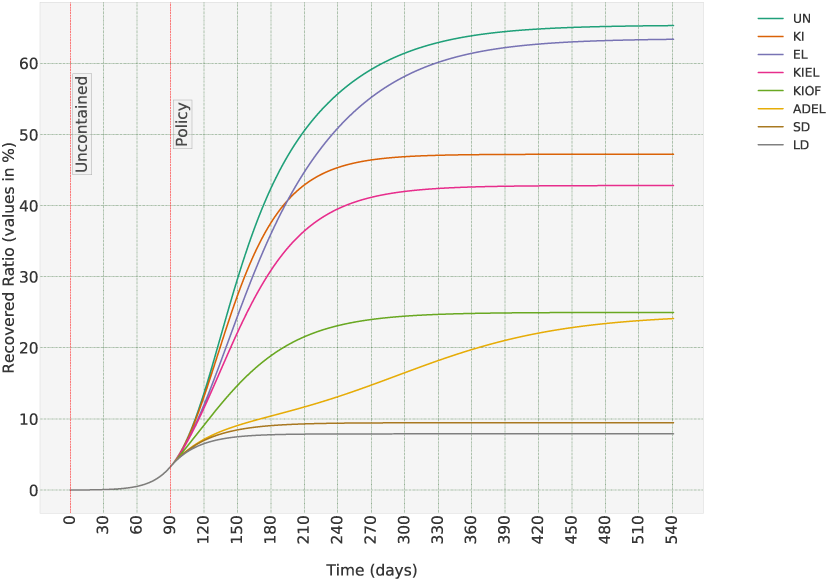
Removed ratios under different policies. Removed population refers to those who were Infectious, and then recovered or died.

One policy that seems to be performing better than expected, is KI, shutting down all schools and universities which is considered as the decrease of contact rates of age groups 0-10 and 10-20 to 10% of the normal times. This policy will decrease the eventual Removed ratio (which is the ratio of the population which has been Infectious at any time during the time period of interest) by more than 20%. To see how this huge decrease has happened, we should have a look at the break-down of the model outputs in age-groups, as shown in Figure 2.3. As can be seen, this policy has decreased the Removed ratios in the first two age groups to almost 0, but its effects tend to decrease for other age groups, to the extent that it is almost non-existent for 70-80 and 80+ age groups. Added to that the fact that 17.4% of the Iranian population is in 0 − 10 age range and 13.9% in 10− 20, we can see why this policy has decreased aggregate numbers of the Removed compartment so much. But we can also see that this policy has less and less effect on the ratio of Removed compartment in older age-groups who are more vulnerable to COVID-19. Hence this policy will have a minimal effect on the mortality rate and the number of patients that need support care in hospitals. Interestingly enough, the first policy that the Iranian government imposed was shutting down schools and universities. At the time of writing this text, offices and companies are ordered to continue their normal operations, after a short break during the Persian new year holidays, while schools are kept closed, and while there is no official decree to keep the elderly at home. A policy that the model show is quite ineffective.

**Figure 2.3:**
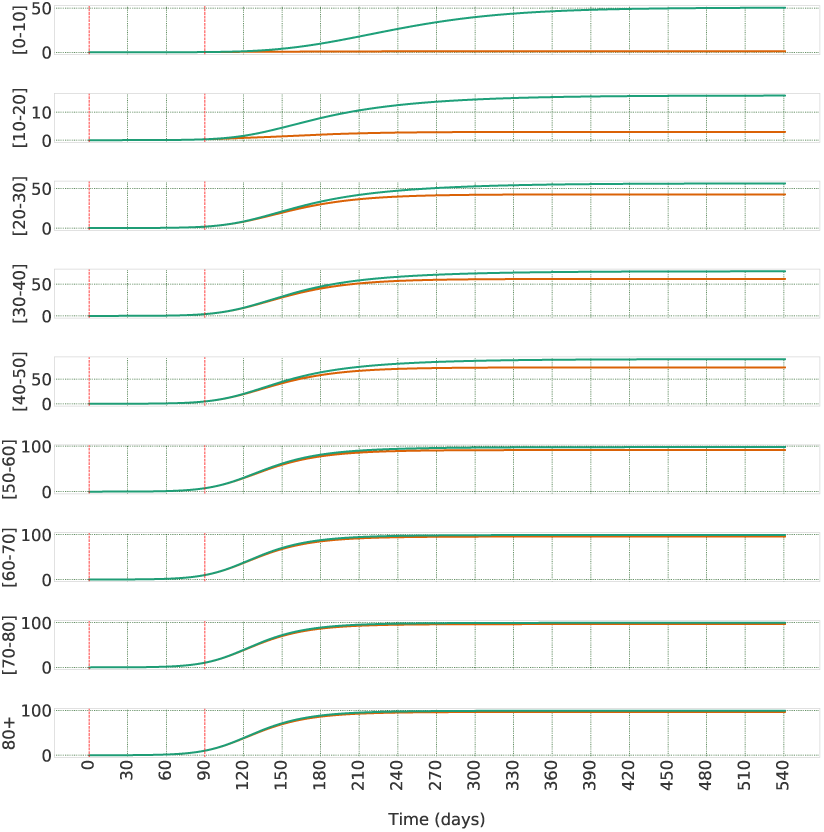
Break-down of the Infectious ratio of each age-group in Iran when the containment strategy includes only shutting down schools and universities. As can be seen, the Infectious ratio in age-groups more vulnerable to disease has not changed significantly.

Table 2 also shows the basic reproduction number, *R*_0_, for each policies. As can be seen in Figure 2.1, and as it is well-known in epidemiology, when *R*_0_ < 1, the disease starts to disappear from the population. But as importantly, bringing the ratio of the Infectious population down to a small enough ratio of the total population can take months, even if we impose a total lock-down strategy (the exact time depends on the ratio of Infectious when we start the policy). And maintaining such a strategy might not be feasible in many countries with a more fragile economy. In the next section, we will see what happens if we switch between different strategies as a long-term plan.

### 3.3 Mitigation Strategies and Long-term Plans

In the previous section, we saw how different containment strategies can affect the spread of the SARS-CoV-2 virus in the population. And we saw that the total lock-down is a necessary strategy when the number of Infectious people grows fast and we need to immediately lower the growth rate. Without such policies, Iran, or any other society for that matter, will face a humanitarian disaster, and the whole purpose of this report is to highlight this imminent threat.

But let’s assume that there is an immediate containment policy in place and the ratio of the Infectious population starts to drop. As we saw, bringing down the ratio of Infectious to a level that can be considered small enough for eradicating the disease from the population can take months. And the economy might collapse if such a stringent strategy imposed for such a time-period. Apart from that, even if the virus is eradicated in one country, in our globalised and connected world there is always the possibility that the virus is re-introduced from other countries who have not imposed containment strategies. So, the natural question is, what policy is suitable as a long-term strategy?

Let’s first see what kind of policy can keep *R*_0_ close to 1.0, so the Infectious ratio does not increase, even if it does not decrease in the short-term. To find the answer to that, we need to run an optimisation scheme to find the coefficients for contact rates in each age group which would bring *R*_0_ from the original estimated 2.28 for the spread of COVID-19 in an uncontained population, to around 1.00. In the simplest case, when we decide to impose a uniform policy to all age groups, the coefficient is 0.4386 or 43.86%. It means that if everybody brings down their direct contacts with other individuals to 44% of what it was in the normal times in Iran, the ratio of Infectious would stay the same or decrease at a slow pace. Please note that this value applies to the age structure of Iran and is different for other countries. This value is calculated using the same optimisation algorithm as explained in SI Section S.2, but with a different objective function.

Now let’s see how these values change if we want to impose different policies for each age group. To make it realistic, I have assumed a different policy for people under 20, between 20 and 70, as the workforce in the society, and above 70. And let’s assume the minimum feasible coefficient is 10% for all age groups. Running the optimisation scheme again, we get values of [15.76%, 14.39%, 99.26%] for the three age brackets, 0-20, 20-70, and 70+. But bringing down the interactions of the working force to 14% of the normal level while not enforcing any changes to the contacts of the elderly does not seem like a sound policy. Let’s set a lower threshold of 40% to the interactions of the working force and keep it at 10% for the other two age ranges. Also, let’s set an upper limit of 20% on the interactions of 70-80 and 80+ age groups, meaning we ask them to self-isolate until further notice, and an upper limit of 50% for people in 0-20 age-bracket. The optimised coefficients are now [38.12%, 54.81%, 19.98%]. So, under such a policy, if kids and teenagers’ interactions are brought down to around 30%, adults to 61% and elderly to 10%, number of Infectious would stay more or less the same for a long period of time.

To bring this idea one step further, let’s assume we plan to impose a switching policy, i.e. we switch between uncontained and different containment strategies at pre-specified time points. To clarify, let’s assume we impose a lock-down strategy for 30 days, and then remove the regulations for 30 days to ease the economic burden, and continue the switching for a few times. Figures 3.1 and 3.2 shows how the spread of the virus changes. As can be seen, the ratio of Infectious follows a sea-saw pattern and in general decreases. Given that we switch to the uncontained policy, it is no surprise that the decrease does not happen as fast as the continuous lock-down policy. Nonetheless, we have managed to stop the Infectious ratio to reach its potential peak value and ease the burden on the healthcare system in the most critical time.

**Figure 3.1:**
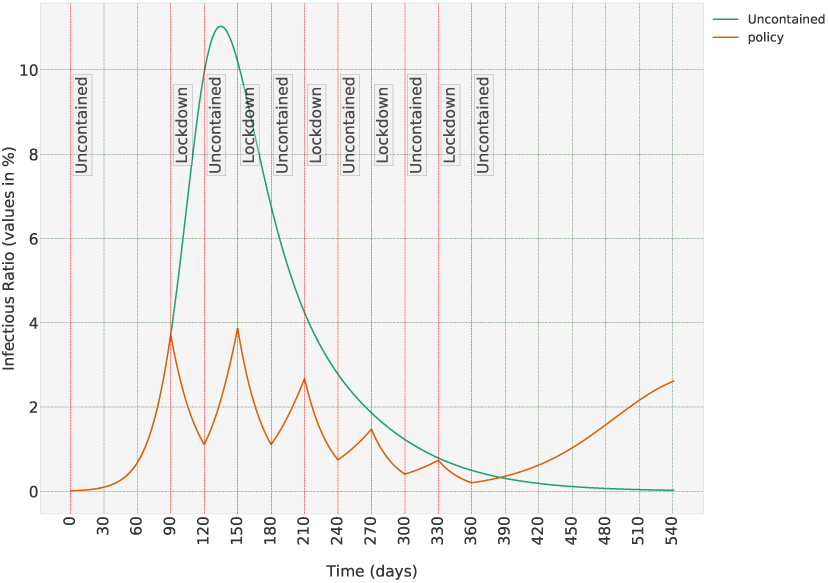
Long-term strategy: switching between uncontained and lock-down scenarios. The Infectious ratio.

**Figure 3.2:**
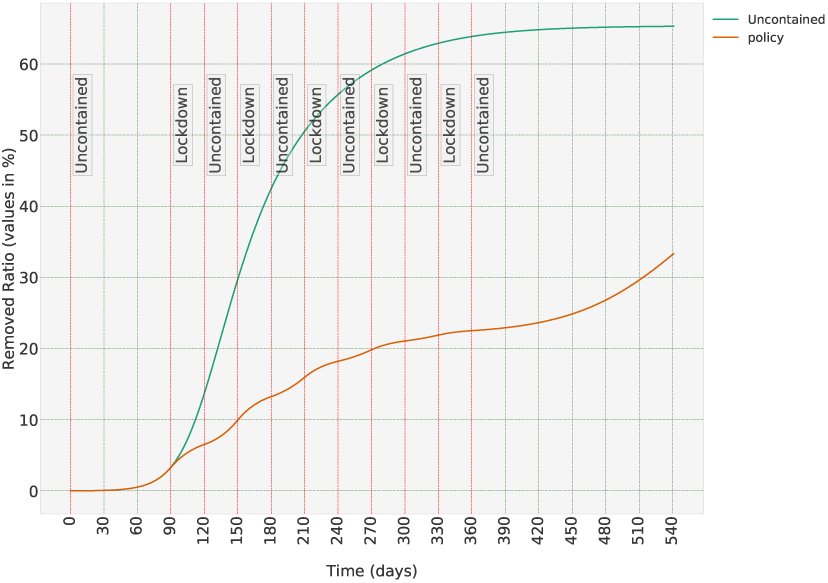
Long-term strategy: switching between uncontained and lock-down scenarios. The Removed ratio.

Now let’s add the *R*_0_ = 1.0 policy to the mix. This time we switch between uncontained, lockdown and *R*_0_ = 1.0 policies. Figures 3.3 and 3.4 show how Infectious and Removed ratios change under such a policy. Since this time there are 60 and not 30 days between uncontained periods, we see a more significant decrease in the Infectious ratio. But that means more people in the population remain susceptible, and if we stop the containment strategy too early, the Infectious ratios bounce back to a greater degree compared to the previous case. And as the last switching policy, let’s consider switching between uncontained and *R*_0_ = 1.0 policies. As expected, such a relatively lax policy has a more attenuated effect on the Infectious ratio. But if any of the previous policies cannot be imposed in a country, policies such as this one can at least ease the burden on the healthcare systems by temporally distributing the Infectious population.

**Figure 3.3:**
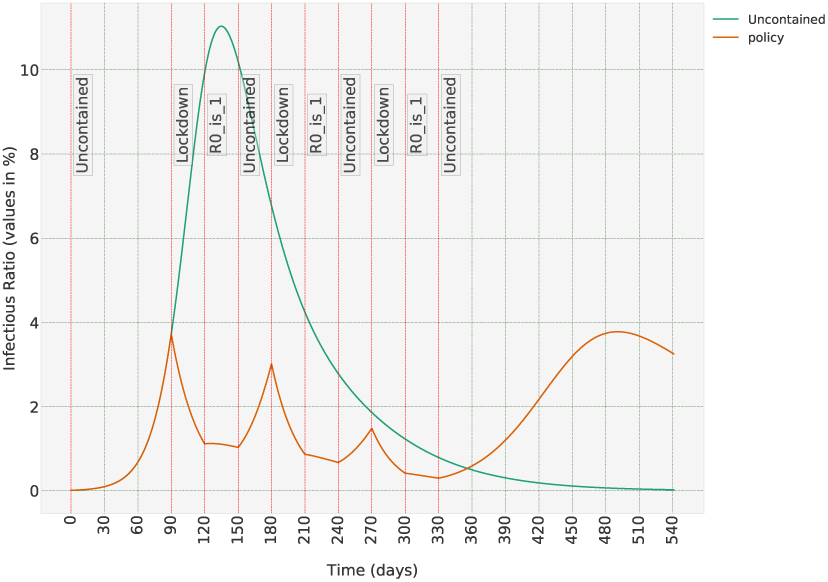
A less strict long-term strategy: switching between uncontained and lock-down and R_0_ = 1.0 scenarios. The Infectious population.

**Figure 3.4:**
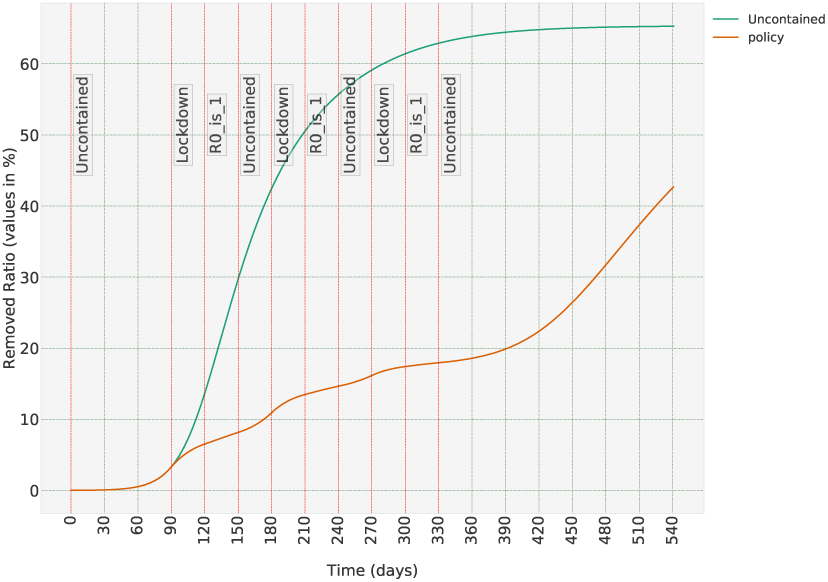
Switching between uncontained and lock-down and R_0_ = 1.0 scenarios. The Removed population.

## 4 Discussions

The method presented in this manuscript can be used to predict the trajectory of the spread of COVID-19 in any population with a known age-structure. The model is capable of predicting the effects of various containment policies imposed on different age groups or on the whole population. The output of the model is the ratio of Infectious and Removed population in each age-group. The Removed population includes those who have been Infectious and then recovered or died. The main advantage of the model is that it needs no a priori knowledge of the interactions between individuals in a population, or how the virus affects each age-group. The contact rates are estimated based on the available data on the spread of COVID-19. The model itself is a set of nonlinear Ordinary Differential Equations (ODEs), hence simulating various containment policies in different time frames does need any special computational power.

But the methodology has some shortcomings that we should keep in mind while interpreting the results. The method assumes the virus affects individuals (of the same age) in different countries similarly. If it is the case that the differences in genetic backgrounds or vaccination histories in different countries can affect the infection rate, such differences would be lost in this model. Also, it is an implicit assumption in the model that the general interactions between people in different countries and societies are in general similar to each other. To clarify the point, if, for example, people older than 70 years in one country live with or have more contacts with the next generations, and in another country, they live in isolation or in nursing homes, then such differences in contacts between age groups are lost in the model. Another issue we should keep in mind relates to the test dataset used in this paper, which was the number of confirmed cases in China in each age-group as of February 11th, and from that, we calculated the desired ratios between the states of the model. And my understanding is that these confirmed COVID-19 cases has been mostly those who have developed symptoms. Hence, the implicit assumption is that the ratio of symptomatic to total number of those infected with SARS-CoV-2 virus is the same for all age groups. If this assumption is proven to be wrong, then the symptomatic to total infected ratio should also be incorporated in calculating the desired ratios between the states of the models.

The method can be extended in various directions to make the results even more useful. We can, for example, divide each age-group into sub-groups based on their vulnerability to the virus, or based on the relative amount of interactions with other individuals in the population. For example, we can divide the population in each group to those with normal levels of contacts, and two sub-groups with one order of magnitude less or more contacts with other members of the populace. Even a rough estimate of the relative numbers of these three sub-groups in each age group can give us more insight into more effective ways to contain the spread of the virus with less social and economic impact. The compartmental model can be extended if necessary. For example, we can add compartment *E* to include latent periods, if it is shown such a time-period is significant in COVID-19. Also, in this manuscript, I have assumed a simultaneous introduction of the virus to different cities and regions in a country, which is not always a realistic choice. Given the fact that nowadays the population structure in different cities/regions in almost every country is known, we can use the model to describe the spread of the virus for each region or city, and then, assuming in and out-flow traffic to each city is known, consider them as exogenous inputs to our system of ODEs. That would allow us to consider time-lags that might exist in the spread of COVID-19 to different parts of a country. But even as it is, this model with its estimated parameters can be a useful tool for different policy-makers in different countries.

## Data Availability

N/A

## Supplementary Material

### S.1 The Mathematical Model

To understand the model discussed in this section, it is enough to know the basics of Ordinary Differential Equations (ODEs) and some basic concepts in linear algebra are needed. Although knowing the following notations and reminding some basic definitions can be helpful.

#### S.1.1 Notations and Some Basic Definitions

ℝ is the field of real numbers and ℝ_+_ is The set of non-negative real numbers. ℝ^*n*^ is The space of column vectors of size *n* of real numbers and ℝ^*n*×*n*^ is The space of *n* × *n* matrices of real numbers. I use *x*_*i*_ to represent The *i*th entry of the vector *x* in ℝ^*n*^, for *i* ∈ {1, …, *n*}. Please note that *x*_0_ is a vector in ℝ^*n*^ that usually represents initial condition. Notation *a*_*ij*_ is used for (*i, j*) entry of the matrix *A. D* = diag (*x*) is an *n* ×*n* diagonal matrix in which *d*_*ii*_ = *x*_*i*_ for all *i. A*^−1^ is The inverse of the matrix *A. I* is the identity matrix of proper dimensions and 0 is the zero matrix of proper dimensions. *σ*(*A*) is the set of all eigenvalues (spectrum) of the matrix *A. ρ* (*A*) is the spectral radius of the matrix *A*, i.e. the maximum of the absolute values of all eigen-values. *µ*(*A*) is The spectral abscissa of the matrix *A*.

*A* ≫*B* means *a*_*ij*_ > *b*_*ij*_, for all *i, j* ∈ {1, …, *n*}. It should not be mistaken with Positive Definite (PD) matrices. *A* > *B* means *a*_*ij*_ ≥ *b*_*ij*_, for all *i, j* ∈ {1, …, *n*} and *A* ≠*B* and *A* ≥ *B* means *a*_*ij*_ ≥ *b*_*ij*_, for all *i, j* ∈ {1, …, *n*}. 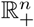 is The positive orthant of ℝ^*n*^, given by {*x* ∈ ℝ^*n*^ : *x* ≥ 0}.

Knowing the following basic definitions can help in understanding the text better.

A matrix *A* is called *Hurwitz*, if *µ*(*A*) < 0.

A real *n* ×*n* matrix *A* = (*a*_*ij*_) is *Metzler* if its off-diagonal entries are non-negative.

The matrix *A* is *irreducible* if and only if for every non-empty proper subset *K* of *N* := {1, …, *n*}, there exists an *i* ∈ *K, j*∈ *N \ K* such that *a*_*ij*_ ≠ 0. When *A* is not irreducible, it is *reducible*.

For any subset 𝒰 of ℝ^*n*^, a point *x*_0_ is called an *interior point* of 𝒰 if there is an open ball around *x*_0_ which is wholly contained in 𝒰. The set of all interior points of 𝒰 is called the interior of 𝒰 and is denoted by int (𝒰).

Consider a continuous-time nonlinear systems of the form:

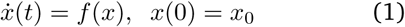

where *f* : 𝒟 ↦ ℝ^*n*^ is a nonlinear vector field on a subset 𝒟 of ℝ^*n*^ and *x*_0_∈ 𝒟 is called the initial condition.

#### S.1.2 SIS Model

Although we only use the SIR model in this manuscript, SIS is also presented here, both in the interest of completeness and to provide a theoretical basis for the SIR discussions. The formulation presented in this section is adopted from [8]. In this model, the population of interest is first divided into two compartments S, Susceptibles, and I, Infectious. For COVID-19, if a latent period exists, it seems to be negligible for practical purposes [12]. Otherwise, we can add another compartment called E, for Exposed, of those who are infected but not yet Infectious. Each compartment is sub-divided into *n* groups. These groups can represent different age groups, different health conditions, professions, etc. In this manuscript, I consider the population in each compartment is divided into *n* = 9 age-groups defined as 0-10, 10-20, …, 70-80 and 80+.

Let *I*_*i*_(*t*) and *S*_*i*_(*t*) be the number of Infectious and Susceptibles at time *t* in group *i* for *i* = 1, …, *n*, respectively. Also, let *N*_*i*_(*t*) = *S*_*i*_(*t*) + *I*_*i*_(*t*) be the total population of group *i*. The total population of each group is assumed to be constant; formally, *N*_*i*_(*t*) = *N*_*i*_. This does not oversimplify the model, especially when the total population is significantly greater than the number of Infectious, which is still the case for COVID-19 at the time of writing this manuscript. But even if that assumption is not deemed realistic for a population, the formulation stated below can be easily altered accordingly.

Here, *β*_*ij*_, the contact rate between groups *i* and *j*, denotes the rate at which Susceptibles in group *i* are infected by Infectious in group *j* for *i, j* = 1, …, *n*. Further, *γ*_*i*_, the transfer rate, is the rate at which an infective individual in the group *i* leaves the Infectious compartment. We also consider birth and death in the population, although to keep the total population constant, we should set the birth and death rates in each age-group to be the same value *µ*_*i*_. Using the mass-action law, the basic SIS model is then described as follows [8]:

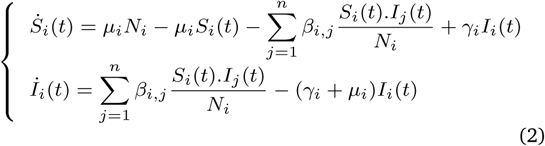

Since the population of each group is constant, it is sufficient to know *I*_*i*_(*t*). If we set *x*_*i*_(*t*) = *I*_*i*_(*t*)/*N*_*i*_ and 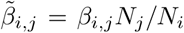 and *α*_*i*_ = *γ*_*i*_ + *µ*_*i*_, we obtain the following differential equation:

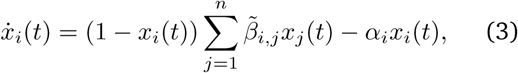

for all *i* = 1, …, *n*. Based on the definition, *x* ∈ *B*_*n*_ where 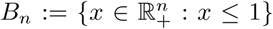. We can write the differential equation (3) in compact form as:

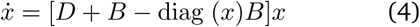

where *D* = − diag (*α*_*i*_) and 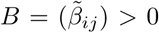. Please note that in the simulations I have assumed the birth and death rate is negligible compared to transfer rate, in other words, *α*_*i*_ = *γ*_*i*_.

The following properties of (4) are easy to check.

i. *f* (*x*) = [*D* + *B* − diag (*x*)*B*]*x* with *D* and *B* defined as above is *C*^1^ in ℝ^*n*^, therefore, the solution for every initial condition in ℝ^*n*^ exists and is unique for all *t* ≥ 0.
ii. The origin is an equilibrium point of (4). This equilibrium is referred to as the disease-free equilibrium (DFE) of the system (4).
iii. System (4) may have an equilibrium in int 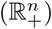 (also referred to as an endemic equilibrium). Conditions for existence of endemic equilibrium for the system (4) depends on parameter *R*_0_, explained below.

One important parameter in mathematical epidemiologically is the *basic reproduction number, R*_0_. There are different definitions for the basic reproduction number. Probably the most common definition is as follows.

##### Definition S.1.1 (Basic reproduction number)

*The basic reproduction number is the expected number of secondary cases produced, in a completely susceptible population, by a typical infective individual during its entire period of Infectiousness* [*7*].

For the SIS model (4), following the reference [8], it can be proved that *R*_0_ = *ρ*(− *D*^−1^*B*). The reproduction number can be used to characterise the existence and stability of the equilibria of (4). As shown in [8, Theorem 2.3], the disease-free equilibrium, i.e. the origin, is a globally asymptotically stable equilibrium of the system (4) if and only if *R*_0_ < 1 (if matrix *B* is irreducible). And the endemic equilibrium, an equilibrium in int 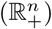, is globally asymptotically stable if and only if *R*_0_ > 1. In other words, the necessary and sufficient condition to eradicate a disease fro a population is to satisfy *R*_0_ < 1 condition.

#### S.1.3 SIR Model

The SIR model is quite similar to SIS, with a minor difference, namely, those who are cured, join the Removed, *R*, population, not *S*. Hence, the formulation for an SIR model is as follows:

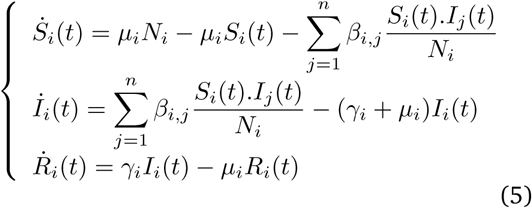

Again, assuming *N*_*i*_(*t*) = *S*_*i*_(*t*) + *I*_*i*_(*t*) + *R*_*i*_(*t*) is constant, similar to what was done in the previous section, if we set *x*_*i*_(*t*) = *I*_*i*_(*t*)/*N*_*i*_ and *y*_*i*_(*t*) = *R*_*i*_(*t*)/*N*_*i*_ and 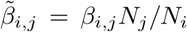 and *α*_*i*_ = *γ*_*i*_ + *µ*_*i*_, *µ*_*i*_ = 0, we obtain the following differential equation:

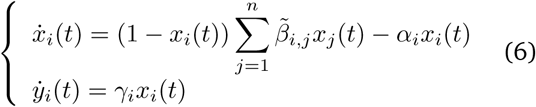

∀*i* = 1, …, *n*. In compact from, (6) can be written as follows:

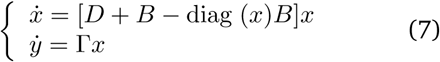

where *D* = − diag (*α*_*i*_) and 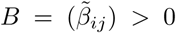 and Γ = diag (*γ*_*i*_) for *i* = 1, …, *n*.

The system (7) has the following properties.

i. *f* (*x*) = [*D* + *B* − diag (*x*)*B*]*x* and *g*(*y*) = Γ*x* with *D, B* and Γ defined as above are *C*^1^ in ℝ^*n*^, therefore, the solution for every initial condition in ℝ^*n*^ exists and is unique for all *t* ≥ 0.
ii. To calculate the equilibria of the system we set *f* (*x*) = 0 and *g*(*x*) = 0. The resulting condition is *x* = 0. Which corresponds to disease-free equilibrium, the origin, and the case that the disease has swept through the population and every remaining person is now either in Susceptible or Remove compartments.
iii. Basic Reproduction Number for (7), can be used using the same formula *R*_0_ = *ρ*(−*D*^−1^*B*).

Property (iii) follows from the discussion in [13, Section 3] and the fact that this equation is derived from the model linearised around the origin. This also means that as the Infectious ratio increases, the effective *R*_0_ becomes less than *ρ* (−*D*^−1^*B*). The observant reader might have noticed that in Figures 3.3 and 3.5, when the policy guarantees *ρ*(− *D*^−1^*B*) = 1.0, the Infectious ratio decreases.

**Figure 3.5:**
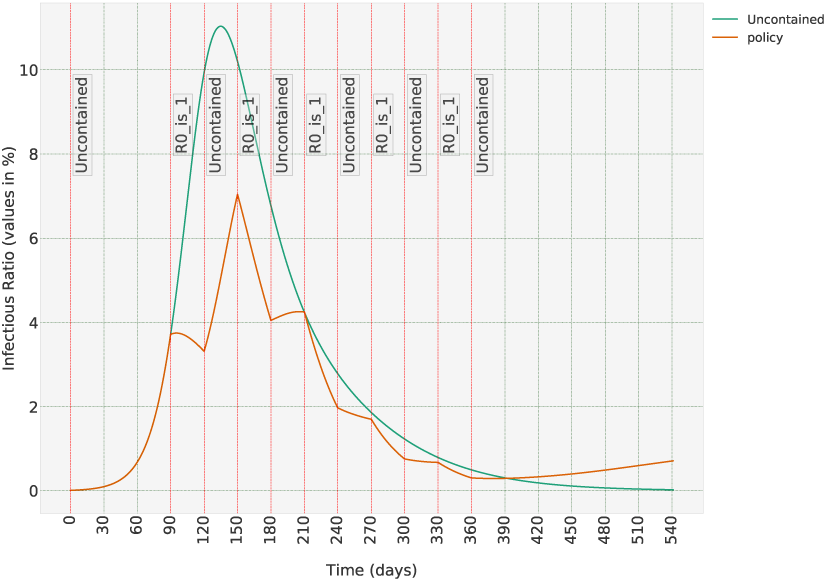
A less stringent long-term strategy when we switch between uncontained and R_0_ = 1.0 scenarios. Unlike the two previous scenarios, the Infectious population grows, but the peak is less than the uncontained case.

**Figure 3.6:**
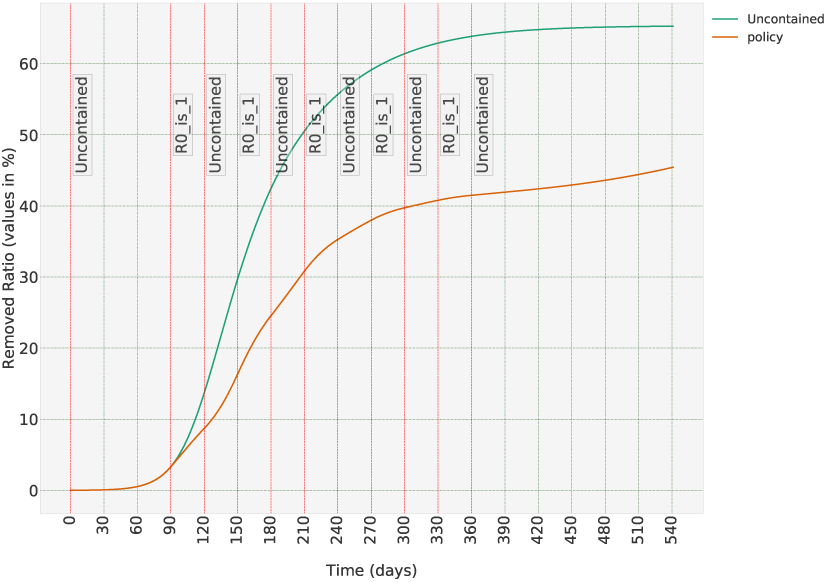
Removed population when we switch between uncontained and R_0_ = 1.0 scenarios.

### S.2 An Optimisation Scheme to Estimate the Parameters of the Epidemiological Model

In order to solve ordinary differential equations (ODEs) (7) or (4), we need to have a reliable estimate of 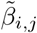 and *γ*_*i*_ for all *i, j. γ*_*i*_ is easy to estimate. If the average duration that individuals in a group *i* are Infectious is 20 days, which seems to be a reasonable estimate for COVID-19 [2], then *γ*_*i*_ = 1/20 = 0.05, given that we have chosen one day to be the unit of time. Estimating 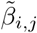, on the other hand, is very difficult, and this section explains how we can estimate contact rates based on real-world data on the spread of COVID-19.

Column C2 in Table 3 shows the distribution of confirmed cases of COVID-19 in different age groups in China as of Feb. 11th [1]. But we should normalise these numbers to the relative distribution of each age group in the general population to be able to compare the differences in how different age groups are affected by the virus. We can do so by dividing values in Column C2 to those of Column C1. We can further divide the resulting numbers to the smallest of them, which happens to be the first row. By doing so, we obtain the last column of Table 3, which shows the normalised relative distribution of infective people in the Chinese population as of Feb. 11th.

**Table 3:**
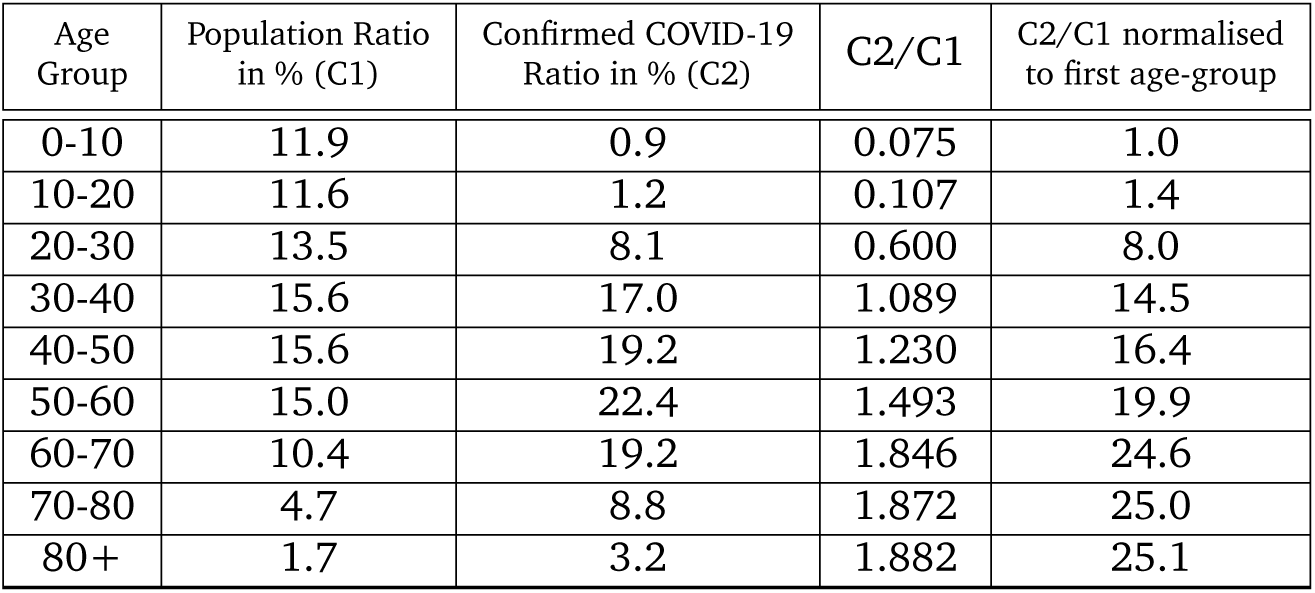
Population distribution in China and the distribution of Confirmed COVID-19 cases in China as of Feb. 11th. To normalise the distribution of confirmed cases, we can divide the values over the population ratio of that age group. The resulting values are then used to estimates parameters of both SIS and SIR epidemiological models.

The optimisation scheme aims to find the contact rates such that at a given time *t*_*g*_, the ratio of the values of the states in Systems (7) or (4) matches the values reported in the last column of Table 3. At the same time, we should keep the basic reproduction number *R*_0_ to be equal to the estimated *R*_0_ for the spread of COVID-19 in an unconstrained population. There are various estimates for *R*_0_ for COVID-19, some are listed in [2]. Most estimates fall in [1.5, 3.5] range. I have chosen *R*_0_ = 2.28, as reported in [15].

Hence, the optimisation problem we need to solve is as follows: finding the matrix *B* such that the following two conditions are satisfied:

i. For a given dialogical matrix *D* and scalar *R*_0_ = 2.28, *R*_0_ = *ρ* (−*D*^−1^*B*)
ii. The relative values of the states of system (4) or (7) at a given time *t*_*g*_ and initial condition *x*_0_ satisfy the values of the last column of Table 3.

But how to choose *t*_*g*_ and *x*_0_? For that, I have relied on reports that the spread of the virus has probably started in a wet market in Wuhan city, in late November [14]. Hence I have set *t*_*g*_ = 75 days (meaning the spread of the virus has initiated 75 days before Feb 11th), and *x*_0_ to be 0 for all groups except 0.0001 in the fourth age group, which corresponds to people aged 40-50. Later on, we will see that the results are robust with respect to the choices of initial conditions and exact value of *t*_*g*_. I have also set constrained for the minimum and maximum of the elements of matrix *B* to be 0.0001 and 0.1 to avoid solutions with anomalous values.

Now that all the required parameters are set, we can solve the optimisation problem to find a *B*_*opt*_. In order to solve the optimisation problem, I have used `sqp` algorithm in `globalsearch` function in `Global Optimisation Toolbox` in Matlab ^c^. Optimisation is done in two steps, in the first step, initial values for matrix *B* are chosen randomly from a uniform distribution. When the optimisation algorithm converges to a solution, the optimisation procedure is repeated, this time with the optimum value obtained in the first step as the initial values. The objective function in the optimisation scheme is the weighted sum of two terms. One is the 2-norm of the difference between the ratio of trajectories of an SIS or SIR model at time *t*_*g*_ with initial condition *x*_0_ with the desired ratio extracted from Table 3. The second term is the difference between *ρ* (−*D*^−1^*B*) and the desired basic reproduction number of *R*_0_ = 2.28. The second term is given a weight big enough so each term is not obscured by the other during the optimisation steps.

The optimisation algorithm runs in 5-15 minutes on 40 hyper-threaded CPUs of type Intel(R) Xeon(R) CPU E5-2687W v4 @ 3.00GHz.

**Note S.2.1** *It should be noted that the values obtained from the optimisation schemes are* 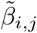 *which are usable only for population distribution in China. But using the relationship* 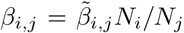, *we can obtain universal values that can be used for all population densities, where N*_*i*_, *N*_*j*_ *are the population ratios in age groups i and j. For each other target population we can use the equation* 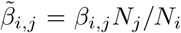 *to calculate the* 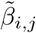 *and then solve ODEs* (4) *or* (7) *to find the spread of the disease in each age group over time*.

**Note S.2.2** *This methodology can be applied to any uncontained or contained population if the effective basic reproduction number, R*_0_ *is known. But given the difficulty in estimating R*_0_ *in a contained population, and the fact that most countries in the world now have a form of containment policy in place, the data collected in early stages of the spread of the virus in China seems to be the best available option as an input to the optimisation scheme*.

#### S.2.1 Sensitivity Analysis

To solve the optimisation problem, I made some assumptions on when and how the disease started to spread in the city of Wuhan. In the absence of concrete facts about the exact moment that the virus was introduced in the human population, we should run a sensitivity analysis to figure out how sensitive is the methodology to the assumptions that were made in regard to the initial conditions.

In order to do so, firstly, we start the value of *t*_*g*_, which was assumed to be 75 days, i.e. I assumed the disease has started to spread 75 days before February 11th. I have changed the value of *t*_*g*_ in [55, 95] days range. For the SIS case, solving the system of ODEs (3), we can calculate *x*(*t*_*g*_) for every *t* − *g* in [55, 95] range, and then calculate the error between the ratios of the states in the resulting vector and the desired values as reported in the last column of Table 3. The maximum relative error over all elements of *x*(*t*_*g*_) and all values of *t*_*g*_ was 0.019. In other words, if the initial guess of *t*_*g*_ = 75 days was wrong and the spread of the disease started any time from 95 days to 55 days before the Feb. 11th date, the values we obtained 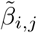 would lead to a ratio between states that match the desired ratios with a maximum error of 0.019 in all age groups. For SIR case, maximum relative error when *t*_*g*_ changes in [55, 95] days range is 0.047.

We can perform similar sensitivity tests on the assumption of the values of initial conditions. As a reminder, to solve the optimisation problem, I assumed *x*_0_ is 0 everywhere except its 4th element (corresponding to the age group 40-50) which is set to `1e-4`. Note that this value represents the ratio of the population of infective in that age group to the total population in that age group. So, if the age group [40-50] includes millions of individuals, which is a reasonable assumption for the regions of China which were reported to be affected by COVID-19 as of Feb 11th. To test the sensitivity to *x*_0_, in the first step, I assumed that instead of [40 − 50] age-group, one other age group among [20,30], [30,40], and [50,60] has an initial condition of `1e-4` while others are 0. For the SIS case, the total relative error over all such cases was 0.051. I then assumed all four age groups from 20 to 60, which are the likely age-groups that might frequent a wet market, have a value of `1e-4` while others are 0, and the maximum error was 0.012, which is even less than the previous case, and I find peculiar. To make the sensitivity analysis even more comprehensive, I have done the same procedure for initial values being of an order of magnitude smaller than what we have assumed so far, it means `1e-5`. Once assuming just one age group in [20,30], [30,40], [40,50], [50,60] has such a value, and then all of them together. When the initial condition was set to `1e-5` for just one of the four age groups between 20 and 60 years, the maximum error in these two cases was 0.050. When the initial condition was set to `1e-5` in all four of them the maximum error becomes 0.008.

Repeating the same procedure in the SIR case, which is the adopted model for COVID-19 in this manuscript, we get different values, but still, the results show small errors. When any of the 3rd, 4th, 5th or 6th elements of *x*_0_ is set to `1e-5` or `1e-4`, maximum relative error in each case for the solution of the SIR system (7) over all four cases is 0.032 and 0.027,respectively. When all four age groups start from those initial values, the maximum relative errors are 0.006, 0.020. To provide a context for what the change of initial condition from `1e-4` to `1e-5` means, we should note that if the initial condition is set to `1e-5` for age group of [40 − 50] years and 0 for other age groups, it takes 69 days until the Infectious ratio in [40 −50] age groups reaches `1e-4`.

To summarise, the sensitivity analysis shows that even if our guess for when and how fast the SARS-CoV-2 virus has started to spread in the human population is off by 20 days, or if the initial number of Infectious is of one order of magnitude lower than what we have guessed, the model obtained from the optimisation scheme generally performs well in reaching the desired values presented in Table 3.

### S.3 Normalised Contact rates for COVID-19

Below, you can find the normalised values of contact rates for the spread of COVID-19 in different populations. These are the results of the optimisation scheme described in SI Section S.2. You can insert these values in the SIR model, (6) in Section S.1.2 or (7) in Section S.1.3, and then solve the ODE with any desired initial conditions to calculate the trajectory of Infectious and Removed populations in each age-group. But please note that in those equations, you need 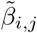 which is calculated as 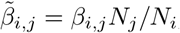, in which *β*_*i,j*_ is the element in row *i*, column *j* of the matrix *B* defined below, and *N*_*i*_ and *N*_*i*_ are the relative ratios of age groups *i* and *j* in the population of interest. As a reminder, the nine age groups used in this manuscript are defined as 0-10, 10-20, …, 60-70, 70-80 and 80+. You can find the age structure of countries in many online resources, for example in [3]. Also, the values of contact rates for both SIS and SIR models, and the code used to run the model and generate the figures we saw in this manuscript can be found in a publicly available online repository [6].

As a last note, when applying a containment policy, the values of the contact rates should be changed accordingly. As an example, when a policy requires the contacts between the first age-group, [0− 10], is decreased to 10% of the uncontained case, the values of the first row of the matrix *B* should be multiplied by 0.1.

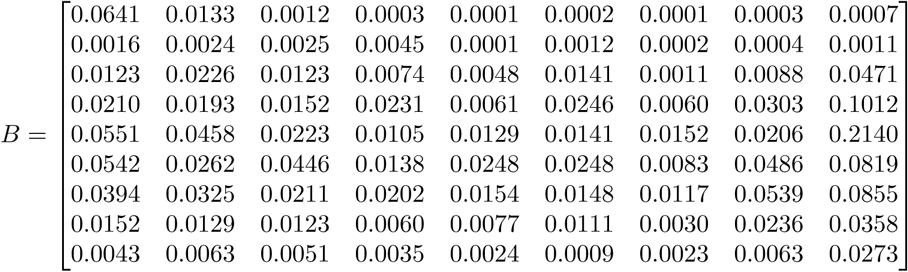

